# Computer Vision Identifies Recurrent and Non-Recurrent Ductal Carcinoma in situ Lesions with Special Emphasis on African American Women

**DOI:** 10.1101/2023.02.08.23285648

**Authors:** Yunus Saatchi, Parker Schanen, Richard A. Cheung, Howard R. Petty

## Abstract

Although the existence of non-recurrent and recurrent forms of ductal carcinoma in situ (DCIS) of the breast are observed, no evidence-based test can make this distinction. This retrospective case-control study used archival DCIS samples stained with anti-phospho-Ser226-GLUT1 (glucose transporter type 1) and anti-phosphofructokinase type L (PFKL) antibodies. Immunofluorescence micrographs were used to create machine learning (ML) models of recurrent and non-recurrent biomarker patterns, which were evaluated in cross-validation studies. Clinical performance was assessed by holdout studies using patients’ whose data were not used in training. Micrographs were stratified by the recurrence probability of each image. Recurrent patients were defined by at least one image with a probability of recurrence ≥98% whereas non-recurrent patients had none. These studies demonstrated no false negatives, identified true positives, and uniquely identified true negatives. Roughly 20% of the microscope fields of recurrent lesions were computationally recurrent. Strong prognostic results were obtained for both Caucasian and African American women. Our machine tool provides the first means to accurately predict recurrent and non-recurrent patient outcomes. We suggest that at least some false positives were true positives that benefitted from surgical intervention. The intracellular locations of phospho-Ser226-GLUT1 and phosphofructokinase type L likely participate in cancer recurrences by accelerating glucose flux, a key feature of the Warburg Effect.

## INTRODUCTION

Human cancer recurrences have not been adequately examined, although cancer recurrences and their sequelae are primary contributors to cancer mortality. Consequently, cancer recurrences have been described as a “black box” (1). Various factors correlate with cancer recurrences including family history, germline mutations such as BRCA1 and 2, immunohistochemistry findings, and gene expression studies (1-4). However, without strongly correlating biomarkers, no definitive, broadly relevant mechanism(s) is available, and without a mechanism, prognostic tests and new drugs are difficult to design. Current FDA-approved tests provide a probability of recurrence, not a definitive prognostic or predictive finding. Instead of relying upon descriptive approaches, moderately correlating factors, or low signal-to-noise methods, we used a hypothetico-deductive approach focusing upon early glycolytic biomarkers that accelerate glucose utilization (5-9). We hypothesize that DCIS recurrences and non-recurrences can be traced to spatial relationships among glycolytic components of cells and tissues. Since Warburg’s seminal work (10), we have known that enhanced glucose utilization accompanies aggressive cancer. Numerous laboratory and animal studies confirm the role of glucose management in cancer (11). Glucose utilization in human cancer is illustrated by the ability of fluorodeoxyglucose and positron emission tomography to visualize aggressive cancers (12). Physical and chemical changes in early rate-limiting glycolytic transporters and enzymes may accelerate glucose uptake and identify pre-invasive lesions competent to become aggressive cancers. To test this concept, we used machine learning (ML) to compare features of glucose flux-controlling steps within DCIS lesions of patients who will or will not experience a cancer recurrence. We found that phospho-Ser226-GLUT1, whose V_max_ dramatically increases after phosphorylation (13), traffics to the ductal epithelial cell periphery in lesions of patients who will experience a recurrence (5). The enzymes PFKL and phosphofructokinase/fructose-2,6-bisphosphatase type 4 (PFKFB4) have been localized to nucleoli (14). We confirmed this finding in ductal epithelial cells of patients who will not exhibit a recurrence (6). In patients who will report a recurrence, PFKL and PFKFB4 are desequestered from nucleoli to accumulate near plasma membranes (6). Thus, peri-membrane regions of ductal epithelial cells of patients who will recur are endowed with special glycolytic advantages. The heightened V_max_ of phospho-Ser226-GLUT1 accelerates glucose equilibration across plasma membranes. PFKL also enhances glycolytic flux due to its diminished feedback regulation. Phospho-Ser226-GLUT1 and PFKL trafficking to membranes serves multiple functions (7). For example, accumulation of glycolytic, pentose phosphate pathway, and glutathione synthesis enzymes near cell surfaces allows substrates to directly enter these pathways. Aggregation of biochemical pathway components accelerates product formation. Castellana et al. (15) have shown that two-step pathways are accelerated 6-fold whereas three-step pathways are accelerated 110-fold by co-clustering. This study leverages these concepts to create ML models of glycolytic biomarker features to predict DCIS patient outcomes. Racial disparities in cancer care are well-known (16); for example, more African American women die from breast cancer after DCIS than Caucasian, Hispanic or Asian women (17). For this reason, we sought to create broadly applicable prognostic machine classifiers exhibiting high performance for African American women.

## METHODS

### Study Design

We test the hypothesis that cancer recurrences and non-recurrences can be predicted using ML. DCIS lesions from recurrent and non-recurrent patients were labeled with biomarkers associated with heightened glucose fluxes. These micrographs were used to train computers. We sought to obtain broadly generalizable computer models to ensure the faithful evaluation of patients. Many patients had full or partial mastectomies, and therefore false positives are expected because surgeries reduce patient recurrences. This limitation is acceptable because it is the only ethical means of extracting statistically significant data.

### Patients and Patient Samples

#### Patient Characteristics

This study included DCIS samples from 185 women who were followed for at least 10 years. At the time of diagnosis, the patients were aged 33-92 years. For recurrent patients, the mean disease-free period to recurrence was 76±62 months. We define a recurrence as any subsequent event, including invasive and metastatic disease, chest wall recurrences, contralateral recurrences, and new DCIS. Data from 70 patients were used for computer training whereas the remaining 115 patient samples were used for holdout studies.

Patients with a “pure” DCIS diagnosis were admitted to this study. For inclusion, certain metadata were required (age, race, non-recurrent/recurrent status, type of recurrence, time to recurrence, surgery, chemo-/radiotherapy, surgical margins, cancer history, relevant family history). All DCIS samples with evidence of lymph node involvement or invasive cancer were excluded from this study. Patients had no previous or concurrent cancer. Tissue samples were obtained from women who had undergone partial mastectomies (67%), total mastectomies (31%), or biopsy alone-no other surgery (1%).

#### FFPE Samples

Archival formalin fixed paraffin embedded (FFPE) pathology samples were obtained from the St. Louis Breast Tissue Registry at Washington University School of Medicine (St. Louis, MO) and Beaumont Hospital (Royal Oak, MI). This tissue sourcing strategy was used to ensure that laboratory personnel did not have access to electronic medical records of patients whose samples were under study. All experiments were blinded or double-blinded and all outcome predictions were made by the software, not the observer (see below). The use of human material was in accordance with the Declaration of Helsinki on the use of material in scientific research. All experiments were approved by the University of Michigan Institutional Review Board (No. HUM000044189).

#### Patient Number

The patient number necessary to obtain the required sensitivity was estimated as described by Jones et al. (18) according to the equation:

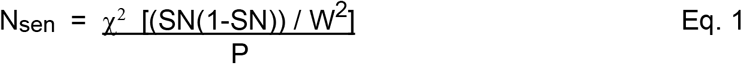

where N_sen_ = the number of patients, SN = the anticipated sensitivity, W = the maximal acceptable width of the 95% confidence interval (0.1) (19), χ_2_ = 3.8, and P = prevalence. Given the distributions below, this is a conservative estimate of the maximal acceptable width of the confidence interval. The anticipated sensitivity is 0.95. As 51/115 holdout patients experienced recurrences in this dataset, we used a recurrence prevalence of 44.3%. This is similar to the abundance found for DCIS patients treated with biopsy alone-no other surgery (46.6±21%), which was obtained by combining 10 published studies (20-29). We found that 41 patients are needed to analyze sensitivity. The specificity was evaluated using:

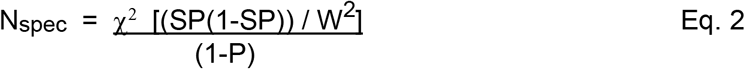

where N_spec_ = the patient number necessary to reach the desired specificity and SP = specificity (0.90). This led to an estimate of 61 patients.

### Immunofluorescence Microscopy

FFPE samples were cut into 5 µm sections. Sections were de-paraffinized and re-hydrated by sequential incubation in a graded ethanol series. After rehydration in PBS with 0.02% Triton X-100 (Thermo-Fisher Sci., Waltham, MA), heat-mediated antigen retrieval in 10 mM citric acid buffer (pH 6.0) was performed on sections. Sections were then blocked using a blocking solution (10% dried milk in PBS) for 1 hr. at room temperature. After blocking procedures, sections were incubated with 2 µg/mL of primary antibodies, rabbit anti-phospho-Ser226-GLUT1 (Millipore-Sigma, cat. no. ABN991), mouse anti-PFKL (Santa Cruz, cat. no. sc-393713) and/or anti-H2A.X (Abcam, cat. no. ab229914) diluted in 1% BSA in PBS overnight at 4°C. After incubation, the sections were washed with PBS. Finally, the sections were incubated with fluorescent second step antibodies - donkey anti-rabbit IgG (cat. no. A10042) and goat anti-mouse IgG (cat. no. A11029) for 1 hr., washed with PBS, and then mounted in Prolong Diamond Antifade medium (Thermo-Fisher Sci.) (5-7).

Fluorescence microscopy was performed as described (5-7). Images were obtained with a Nikon TE2000 microscope (Nikon, Melville, NY) with a 20x (0.50 NA) objective, 1.5x optovar and a back-illuminated Andor iXon electron-multiplying charge-coupled device camera (Model DV-888; Andor Technology, Belfast, Northern Ireland). The 20x objective with or without a 1.5x optovar was used to permit the capture of micrographs containing information from the duct and the stroma to support prognostic analyses, as too high a magnification could miss features correlating with recurrences. A moderate NA was chosen to provide the highest possible resolution while avoiding objective-mediated optical sectioning of the tissue section, which would lead to a loss of information. Images were captured and processed with MetaMorph software (Molecular Devices, San Jose, CA). To reduce shot noise, each micrograph was an average of 10-15 separate image acquisitions, with each image acquired for 0.2 sec. To reduce read noise, the EMCCD chip was cooled to -95° C. Typical camera settings were multiplication gain, 100; vertical shift speed, 3.04 msec./pixel and 14-bit digitization at 10 MHz.

### Computer Procedures

#### Machine training

We employ supervised machine learning to create computer models that recognize biomarker patterns within tissue sections originating from patients who will or will not exhibit subsequent cancer recurrences. Custom Vision is Microsoft’s (Redmond, WA) computer vision application programming interface built atop convoluted neural networks. The software was deployed as a multiclass (tags, recurrent or nonrecurrent) and general domain problem. Micrographs from 70 patients were available for computer training. The computer was separately trained with micrographs of PFKL and phospho-Ser-226-GLUT1-stained tissues (6, 7). Prior to model building, micrographs were triaged. Micrographs with poor focus were excluded. Images containing artifacts such as tissue section folds, fluorescent label trapping or other artifacts were not used in training. Sections that did not contain enough micrographs and/or ducts were discarded. As peripheral biomarker labeling patterns were often observed for patients’ samples prior to cancer recurrences, but less frequently in nonrecurrent samples, machine training was based on tissues exhibiting peripheral or non-peripheral labeling patterns of these two glycolytic biomarkers. Thus, to train the computer to recognize recurrent images, glycolytic biomarker clustering at the cell periphery of micrographs from recurrent patient samples were used. On the other hand, to select images for non-recurrent training, micrographs without peripheral biomarker clusters of non-recurrent samples were selected whereas images from non-recurrent patients with peripheral biomarker clustering were not used in training. This approach minimizes confounding errors by limiting the contribution of presumed false negatives (recurrent patients without peripheral labeling) and false positives (non-recurrence patients with peripheral protein labeling) from the training data. This approach is justified by highly significant findings (P < 0.0001; see below).

#### Cross-Validation Studies

Computer model performance was assessed by calculating precision and recall using 5-fold cross-over analyses. The precision is:

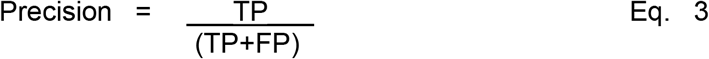

where TP indicates true positives and FP indicates false positives. The recall was calculated according to Eq. 4:

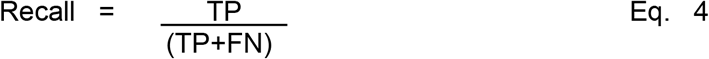

where FN indicates false negatives. Data were evaluated using precision-recall curves, which plot these variables across threshold values. Precision-recall curves are much less sensitive to differences in the numbers of patients in each group than receiver operating characteristic plots (30).

#### Overfitting

Overfitting is a serious issue in medical machine learning. Our protocol was designed to minimize overfitting of data. The application programming interface does not permit duplicate images, thereby reducing the potential for overfitting. As the Azure platform uses low-rank filters for image classification with convoluted neural networks, computationally efficient models that avoid overfitting and generalize better are obtained (31). Operationally, the possibility of overfitting is minimized by using additional patient samples and by collecting multiple micrographs per section. Image augmentation may also reduce overfitting by increasing the data set size. By using fluorescence microscopy, which has a high signal-to-noise ratio, we reduce the possibility of overfitting noise. As stroma and ductal regions of tissue are important in predicting recurrences (7-9), there is no extraneous image information to distract the classifier. Very high levels of precision and recall are signs of overfitting, and the model should be discarded. Overfitting was also avoided by limiting training time; thus, smaller data sets require less time than larger data sets. Also, for each dataset, the performance of the ML model in classifying holdout data was noted for different training times.

#### Data set tuning to improve recurrence predictions for African American women

In preliminary studies, we found that the classifiers did not perform well using micrographs obtained from African American DCIS tissue lesions. To address this concern, micrographs from Caucasian and African American patients were evaluated by direct inspection. After balancing the recurrent and non-recurrent data sets for blood vessel size and for lipid droplet content, we found that the performance of holdout samples of Caucasian and African American women was indistinguishable (see below).

#### Computed outcome prediction

Using our ML models, we compute well-calibrated cancer recurrence probabilities to stratify patient outcomes. We tested a verification (holdout) data set containing micrographs of each patient (N=115) using the Custom Vision application. In general, we sought to obtain roughly 25-50 micrographs per biomarker from each patient for computational analysis. No patients in the holdout data set were included in the training data set. The cut-point was set at a 98% recurrence prediction. In most cases, the computed probability of outcome was 0-3% for individual micrographs from non-recurrent patients and 98-100% computed probability of recurrence for micrographs deemed to be recurrent.

### Statistical Tests of Outcomes for Verification Studies

Computed predictions were statistically evaluated. Two-by-two contingency tables were constructed using clinical patient outcomes and computed outcomes. Fisher’s exact test was chosen because there is a small value in the FN cell. Test accuracy is:

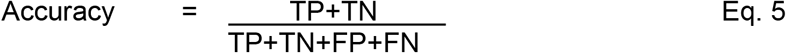

Another means to evaluate classifier performance is the F1 score. The F1 score is the harmonic mean of the precision and recall (Eq. 6).

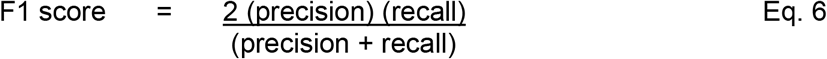

Both accuracy and F1 score are widely used in ML studies.

We also employed the Matthews correlation coefficient (MCC) to evaluate the test (32) because it is not affected by unbalanced data sets. It is calculated as:

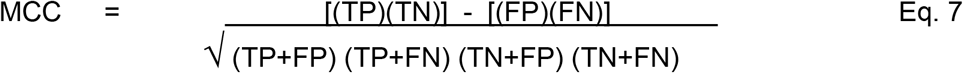

### Estimating Recurrence Parameters

As noted above, DCIS recurrences among patients treated with biopsy alone, Rec_biopsy_, is 47±21% (range 14-73%) (20-29). As most patients undergo full or partial mastectomies, we estimate the impact of these procedures on patient outcomes. We calculate test performance using two conditions: 1) we determine performance metrics assuming surgery has no effect on patient outcomes and 2) we evaluate test performance assuming surgery causes FPs due to the removal of cancerous tissue thus converting TPs to FPs. The impact of surgery on test performance is illustrated in Fig. 1. As described above, we estimate that ∼47% of the patients will recur; the remainder will not recur. There are successful surgeries that are computationally recurrent, but clinically non-recurrent due to surgery; these will appear in data as FP. Using NRec_biopsy_ and the observable recurrence rate after partial mastectomies (NRec_partial_) of 10% (33), we estimate the impact of surgery (N_surgery_) as:

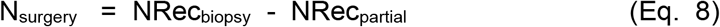

**Fig. 1.**
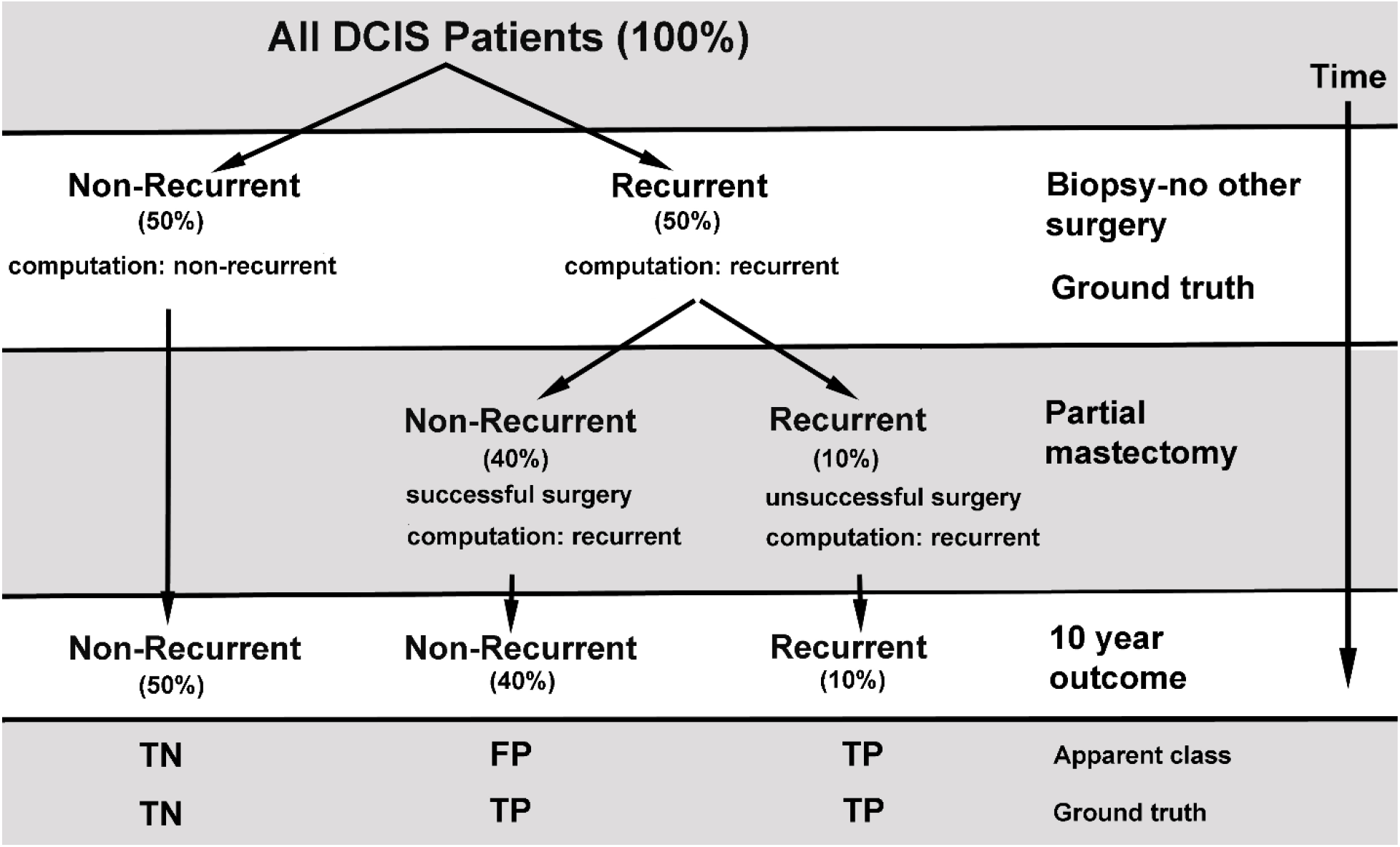
The Effect of Mastectomies on Observed Cancer Recurrence and Non-Recurrence Rates. Up to 50% of the patients diagnosed with DCIS, but not treated with full or partial mastectomies, will recur; the remainder will not recur (20-29). After partial mastectomy, roughly 10% of the population will recur (TP) (33). This suggests that many patients who were computationally recurrent (with relevant changes to the glycolytic apparati) with non-recurrent outcomes became non-recurrent due to surgical intervention (FP). Thus, ∼10% of patients treated with partial mastectomies will recur (TP), and the remainder is computationally positive but clinically non-recurrent (FP) or computationally negative and clinically non-recurrent patients (TN).

We use N_surgery_ to correct confusion matrices for the effect of surgeries on patient outcomes, which is ∼37%.

## RESULTS

### Cross-Validation Studies of ML Models

To verify the performance of phospho-Ser226-GLUT1 and PFKL ML models, we assessed their precision and recall in cross-validation studies. Training data sets were randomly divided into five subsets. In these five-fold cross-validation studies, computer training used 80% of the micrographs for each biomarker, and then the model was tested using the remaining 20% of the micrographs for each of the five subsets. The performances of ML models are illustrated using precision-recall curves (Fig. 3A, B). Both classifiers exhibit excellent performance in cross validation studies and are used to predict patient outcomes in holdout studies.

**Fig. 2.**
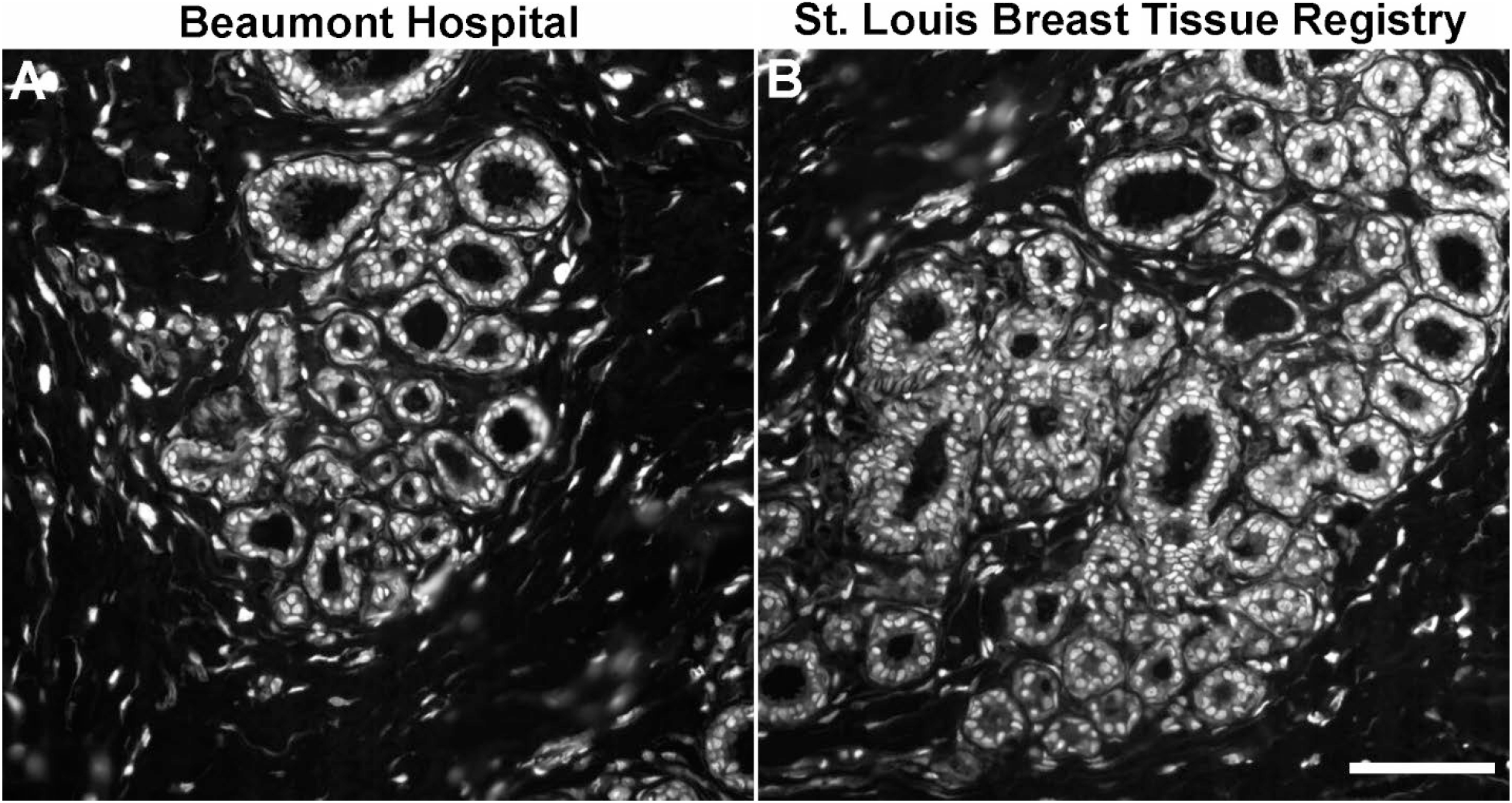
Comparison of Tissue Labeling Using Different Sources of DCIS Tissue. Tissue sections of DCIS lesions were prepared then stained with anti-phospho-Ser226-GLUT1 and then with a fluorescent second-step antibody. Both panels show non-recurrent tissue sections. Panel A shows a micrograph of an image derived from a sample from Beaumont Hospital. Panel B shows a similar micrograph of a sample from the St. Louis Breast Tissue Registry. There is no discernable difference in the appearance of the images. (Bar=200µm).

**Fig. 3.**
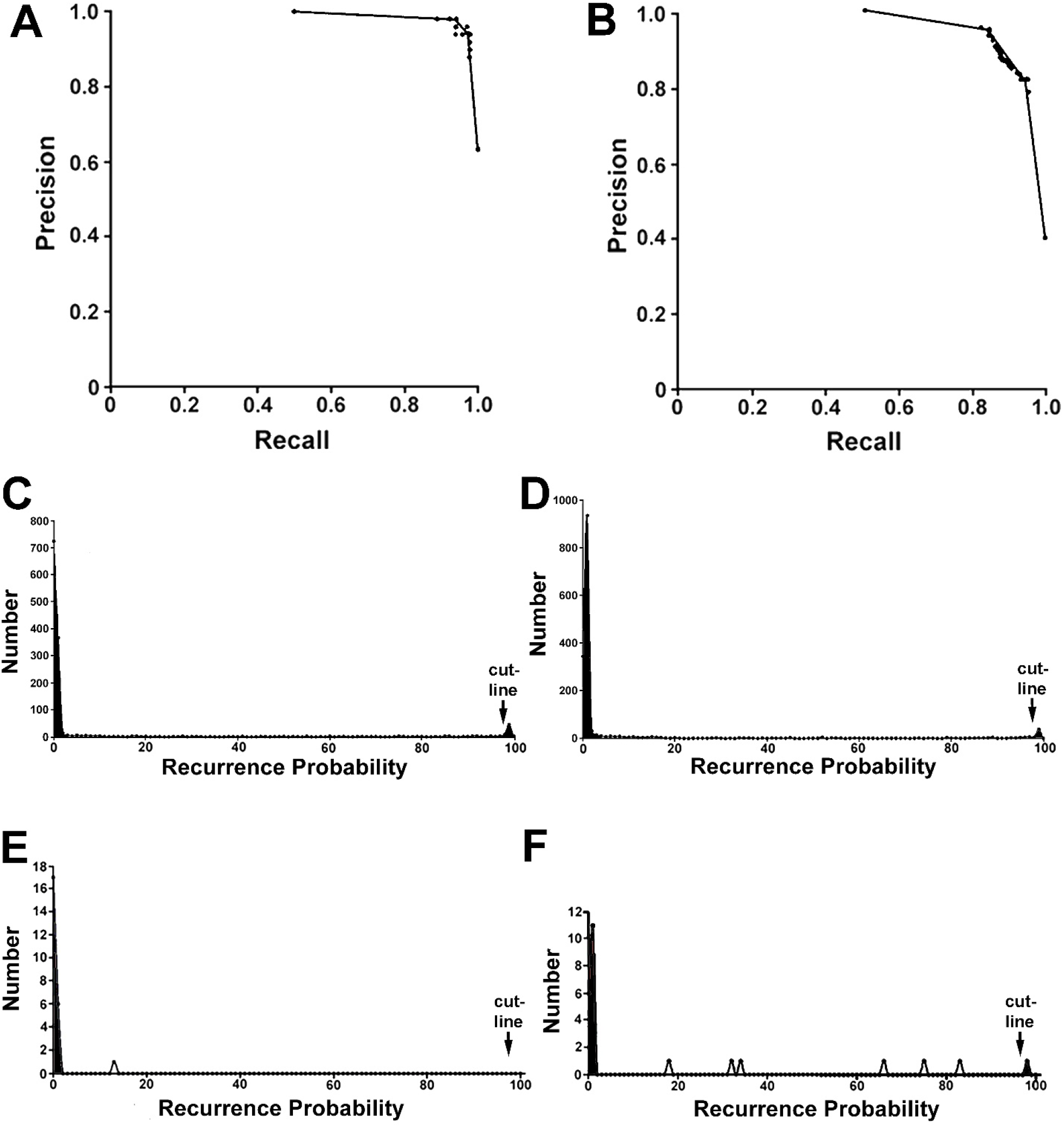
Computer Findings and Prognoses. Panels A and B show the performances of the phospho-Ser226-GLUT1 and PFKL classifiers. In these cross-validation studies, the training data sets were randomly divided into five subsets. The computer was trained using 80% of the micrographs for each biomarker and outcome, and then evaluated against the remaining 20%. This process was repeated for all subsets to estimate an overall precision (TP/(TP+FP)) and recall (TP/(TP+FN)) of each classifier’s training data set. Panel A shows the precision-recall curve of the phospho-Ser226-GLUT1 model. Panel B shows the precision-recall curve of the PFKL model. The phospho-Ser226-GLUT1 model showed an AUC (area under curve) of 0.96 whereas that of PFKL is 0.86. Both classifiers showed excellent performance on individual micrographs. Panels C and D show the summed distributions of recurrence predictions for holdout patients (number of micrographs at ordinate and probability of recurrence at abscissa). C) This study uses ML models to assess the probability that a microscopic field originates from a recurrent patient. This panel plots data from DCIS microscope fields of phospho-Ser226-GLUT1-labeled tissue sections for all patient holdout micrographs. The number of micrographs at each % probability is plotted against the % probability of recurrence. The separation of the non-recurrent and recurrent mode values along the x-axis is very large. The mode values also demonstrate a high signal-to-noise ratio. Substantial improvements in sample analyses compared to conventional tools are obtained. D) Parallel studies of PFKL-labeled DCIS tissue sections are shown. Panels E and F show the distributions of recurrence predictions for a non-recurrent patient and a recurrent patient, respectively (number of micrographs at ordinate and probability of recurrence at abscissa; note that the range of the ordinate in these graphs is much smaller than panels C and D). E) The computed probabilities of recurrence determined by machine analysis of micrographs from a clinical and computed non-recurrent patient are shown. All image recurrence probabilities are low (<15%). F) In contrast to panel E, quantitative data for a clinical and computed recurrent patient sample is shown. In addition to the spike at low probabilities, several intermediate probability levels and one high probability of recurrence observation (98% recurrence probability) are apparent. A single microscope field at this level is sufficient for the computed prediction of a recurrence.

### Clinical Verification using Holdout Data

Conventional diagnostic tests often rely upon a single variable, such as a spectrophotometric intensity, to identify a clinical condition. Individuals within healthy or ill populations test at multiple levels, leading to overlapping curves. The overlapping regions cause FN and FP. This is not the case with our machine studies where we calculate recurrence probabilities to stratify samples.

#### FFPE Source Comparison

This study uses two sources of FFPE DCIS tissue samples, the St. Louis Breast Cancer Registry and Beaumont Hospital. To provide support for this strategy, we compared images using these tissue sources. Fig. 1 shows immunofluorescence studies of samples from Beaumont Hospital (panel A) and the St. Louis Breast Cancer Registry (panel B). These non-recurrent tissue samples were labeled with anti-phospho-Ser226-GLUT1. This biomarker was chosen because their internal sequestration is easily observed in samples from patients who will not recur. In contrast, GLUT1 is found at the periphery of epithelial cells of all DCIS lesions (5). Phospho-Ser226-GLUT1 is found in the nuclear region of samples from both the St. Louis Breast Cancer Registry and Beaumont Hospital. Additionally, we have previously used the National Disease Research Interchange to obtain DCIS and normal adjacent tissue for labeling (6). In other studies, we have used FFPE tissue samples from Precision Medical, the Cooperative Human Tissue Network, Biochain, and others with this protocol. Samples from the St. Louis Breast Cancer Registry and Beaumont Hospital were used in this study because the associated metadata met our inclusion criteria.

#### Patient Scores and their Distributions

Holdout verification studies were performed to determine the models’ clinical applicability. Patient holdout studies were performed using biopsies from 115 women. We computed the cancer recurrence probability for each micrograph to stratify patient data to predict outcomes. The probability that a micrograph was linked to a cancer recurrence follows a bimodal distribution for phospho-Ser226-GLUT1 and PFKL (Fig. 3C, D). The probability that a micrograph originated from a recurrent lesion was often high (∼98-100%) or low (0-2%). Fig. 3C shows a recurrence probability plot for phospho-Ser226-GLUT1 labeled micrographs vs. the number of micrographs at each probability. Recurrence probability plots (panels C and D) quantitatively illustrate lesion heterogeneity. Roughly 20% of the microscope fields of recurrent lesions are computationally recurrent, whereas the remaining images are non-recurrent. As the DCIS population data of Fig. 3C show, the recurrent group mode value is 47 and the background level is 1.15 ± 1.20 (mean ± SD). The mode is 37 standard deviations above background noise, indicating a strong ability to detect cancer recurrences.

To complement phospho-Ser-226-GLUT1 assessments, we used PFKL, which undergoes spatial changes in DCIS lesions destined to recur (6). Using a second ML model, the population distribution of individual PFKL scores for DCIS patients was obtained (Fig. 3D), which resembles that of phospho-Ser226-GLUT1 (panel C). Recurrence determinations require that one or more micrographs of phospho-Ser-226-GLUT1 or PFKL have a ≥98% recurrence probability.

In addition to providing information about all patients, we also show individual patient score distributions in Fig. 3E and F to demonstrate clinical utility. Panel E shows a TN (clinically non-recurrent and computationally negative) and panel F shows a TP (clinically recurrent and computationally positive); note the presence of a microscope field at a 98% recurrence probability in panel F. To visualize these samples, Fig. 4 provides fluorescent micrographs of DCIS lesions labeled with anti-phospho-Ser226-GLUT1 that were scored as TP, FP, and TN by the ML model. Accumulation of proteins at cell peripheries is an important indicator of recurrent cells (5-8). However, this is neither a necessary nor sufficient condition for a recurrent prediction; accumulation at the cell periphery correctly predicts about 70% of the patient outcomes (5, 6) (see also following sub-section). It should be noted that FP micrographs are very similar to TP micrographs, but not TN micrographs.

**Fig. 4.**
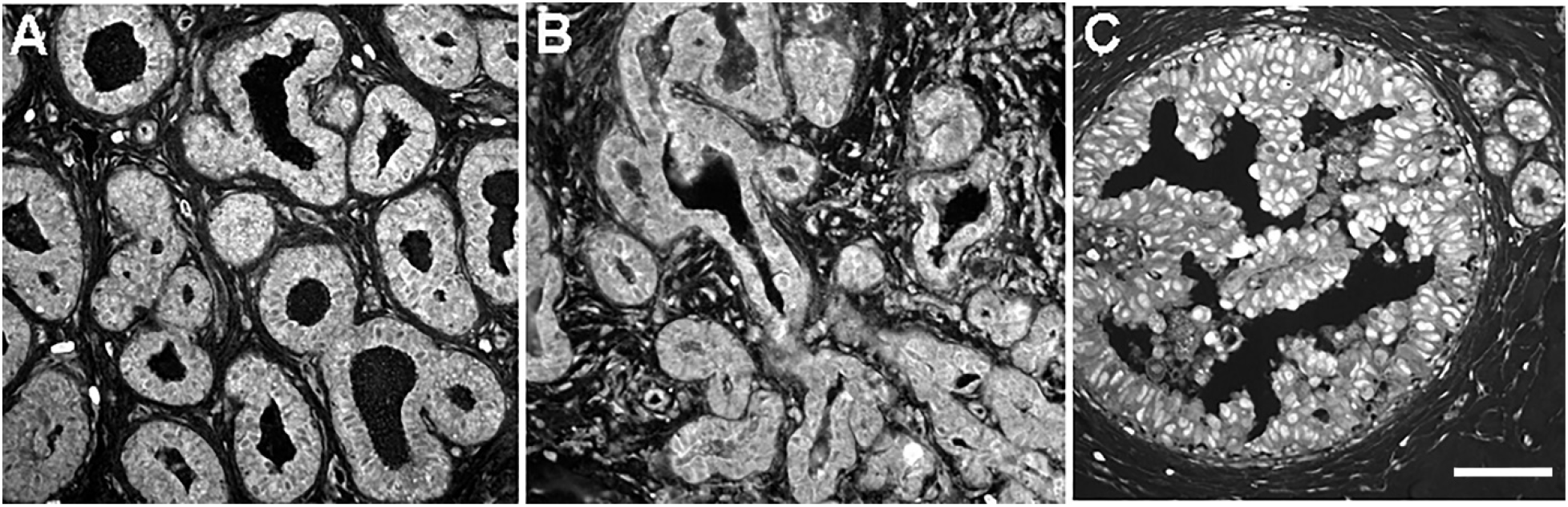
Representative Micrographs of True Positive, False Positive, and True Negative Samples. Tissue sections of DCIS lesions were prepared then stained with anti-phospho-Ser226-GLUT1 and then with a fluorescent second-step antibody. Panel A shows a true positive sample (clinical recurrence, computed recurrence). Panel B shows a false positive micrograph (clinical non-recurrence, computed recurrence). Panel C shows a true negative micrograph (clinical non-recurrence, computed non-recurrence). From observation alone, the false positive is much more akin to the recurrent than the non-recurrent sample. (Bar=200µm).

A primary factor in outcome predictions was the computed status of phospho-Ser226-GLUT1. Most of the recurrent class projections for TP and FP samples were made with a single positive biomarker (e.g., either phospho-Ser226-GLUT1 or PFKL) and 30-40% exhibited positive computational results for both biomarkers. However, PFKL also impacted the recurrent outcome predictions for those cases in which phospho-Ser226-GLUT1 computations were negative. PFKL recurrent machine classifications were required for 4/115 TP predictions. PFKL recurrence identifications reduced the total number of FN by 6/115 women. PFKL trafficking is an important contributor to prognostic calculations because: it is a strong indicator of patient outcome (6) and the reduction of FN is important for clinical use.

#### Non-Canonical Tissue Patterns

In this and previous studies (5-9), we focused upon presenting representative examples of recurrent and non-recurrent tissue labeling patterns. For practitioners, however, it is important to know the rules and the exceptions to the rules. It is therefore important to consider the properties of the entire population, including non-representative examples. To illustrate unconventional findings, a true negative (clinically and computationally non-recurrent) is shown in Supplemental Fig. 1. Supplemental Fig. 1A, B show the nucleolar biomarker histone HA2.x and phospho-Ser226-GLUT1. HA2.X and phospho-Ser226-GLUT1 co-localize in nucleoli of non-recurrent DCIS lesions. Although PFKL is found in nucleoli of non-recurrent DCIS lesions (6) and other tissues (14), this patient displayed peripheral PFKL labeling (Supplemental Fig. 1C), which is observationally consistent with the recurrent phenotype. Computational studies using both phospho-Ser226-GLUT1 and PFKL models predicted that the patient was non-recurrent (> 200 micrographs were tested). Thus, the ML models are more reliable than visual inspection.

#### Samples from African American Women

Preliminary studies showed that the ML models did not perform well for African American women. Therefore, new classifiers were built and “tuned” as described under Methods. To ensure that these new ML models performed well using African American women, the patient holdout group was composed of similar numbers of Caucasian and African American women. The disease-free time to recurrence did not differ significantly (Caucasian 79 ± 65 months and African American 68 ± 53 months (P=0.54). Quantitative data on classifier performance are summarized in Table 1. No FNs were observed. No significant differences in assay performance were noted in comparing all women tested to African American women (e.g., the non-recurrent populations of all women tested and African American women tested demonstrated a P value of 0.55 using Fisher’s exact test). To determine if assay performance varied with surgical procedures, we analyzed clinically non-recurrent patients (FP and TN) treated with either full or partial mastectomies (Table 1). Fisher’s exact test showed that there were no significant differences among these groups (P=0.39).

**Table 1.** Summary. Holdout Experiments^1,2^

| A | B | C | D | E | F | G | H | I | J | K |
| --- | --- | --- | --- | --- | --- | --- | --- | --- | --- | --- |
| Clinical Outcome Post-Mastectomy<sup>1</sup> | Computed Outcome at Time of Surgery | Class | All Women Tested Uncorrected | African-American Women Uncorrected | Women with Unreported Race | All Women After Full Mastectomy Uncorrected | All Women After Partial Mastectomy Uncorrected | Biopsy- No Other Surgery | All Women Corrected | Comments |
| Recurrence | Recurrence | TP | 51 | 11 |  | 16 | 35 | 1 | 79 | highly accurate detection of recurrences |
|  | Non-Recurrence | FN | 0 | 0 |  | 0 | 0 |  | 0 | no ill patients missed |
| Non-Recurrence | Recurrence | FP | 28 | 20 |  | 11 | 12 |  | 0 | outcomes improved by surgery |
|  | Non-Recurrence | TN | 36 | 20 | 4 | 9 | 17 | 1 | 36 | patients do not require intervention |
| Fisher's Exact Test |  |  | <0.0001 | 0.0035 | - | 0.046 | <0.0001 | - | <0.0001 |  |

#### Computed Recurrence Percentages do not Differ for True Positives and False Positives

We considered the possibility that cancer recurrences might be related to the density of recurrence-positive microscope fields (positive microscope fields per unit area), not to the presence of just one recurrence-positive microscope field. In other words, FP could be qualitatively identical to TP, but the density of recurrence positive microscopic fields in FP samples could be too low to successfully support a subsequent recurrence after surgery. To test this idea, the recurrence positive microscope fields and recurrence negative microscope fields were quantified for TP and FP patient samples. The TP and FP distribution curves were skewed positively. The TP samples exhibited 20.2 ± 19.5% (mean ± SD) recurrence positive microscope fields. For FP samples, we found 19.3 ±16.5% recurrence positive microscope fields. A value of P=0.38 was obtained for these data, which does not support the hypothesis that cancer recurrences can be explained by the burden of recurrence positive microscope fields in DCIS lesions.

#### Further Statistical Analyses of Predicted Outcomes

We evaluated several statistical features of holdout data. Data were evaluated with two assumptions: 1) that surgery has no effect on patient outcomes and 2) that surgery causes some FP by removing tissue that would have subsequently recurred. Fisher’s exact test yielded P<0.0001 for both raw and corrected data (Table 1). The F1 score and accuracy for raw data are 0.79 and 0.77, respectively. The Matthews Correlation Coefficient (MCC) is 0.60 while the true positive rate (TPR) is perfect. Raw data were corrected using the estimated value of N_surgery_ = 37% (Fig. 2; Eq. 8), which reduced the number of false positives. The corrected test accuracy and F1 score are 1 and 1, respectively. At a cutline of 98%, there is no overlap between the recurrent and non-recurrent populations, thus, machine analysis constitutes an ideal test. The TP rate is 1 for both raw and corrected data. The MCC and the false positive rate are also improved by corrected data (Table 2). Under both surgical assumptions, a population of TN (clinically non-recurrent and computationally non-recurrent) is identical and accounts for ∼36% of the DCIS patients in these studies. These findings support the prognostic inferences that satisfy the objectives of this study.

**Table 2.** Performance Metrics of Patient Holdout Studies with and without Correction for Surgical Intervention.

| Test | Equation | Surgery has no Effect on Outcomes<sup>1</sup> | Surgery Causes False Positives<sup>2</sup> |
| --- | --- | --- | --- |
| Precision | 3 | 0.65 | 1 |
| Recall | 4 | 1 | 1 |
| Accuracy | 5 | 0.77 | 1 |
| F1 score | 6 | 0.79 | 1 |
| Mathews Correlation Coefficient | 7 | 0.60 | 1 |
| TPR<sup>3</sup> |  | 1 | 1 |
| FPR<sup>3</sup> |  | 0.56 | 1 |

1) Data are from Table 1, column D.
2) These data are the raw data of Table 1, column J corrected for FP caused by surgical intervention, as illustrated by Eq. 9.
3) More conventional metrics of assay performance are the true positive rate (TPR) or sensitivity is $TP/(TP+FN)$ . The false positive rate (FPR) or specificity is $TN/(TN+FP)$ .

## DISCUSSION

DCIS may, or may not, lead to breast cancer recurrences. Roughly, 50% of DCIS patients treated with biopsy alone will later develop recurrent breast cancer (20-29). In the absence of an evidence-based tool to classify DCIS lesions as recurrent or non-recurrent, nearly all patients are treated with surgery (34-38). We now report the first evidence-based tool using broadly generalizable ML models to identify DCIS lesions that do or do not precede recurrences.

An advantage of this machine test is the very low number of false negatives. Immunofluorescence microscopy has a superior signal-to-noise ratio (∼100:1) and spatial resolution in comparison to immunohistochemistry. It also has substantially greater specificity than H&E staining. In contrast to many laboratory tests, these class-conditional probability calculations yield distributions of non-recurrent and recurrent populations that do not overlap (Fig. 3C-F), presumably due to the clear ability to use all the spatial biomarker information inside and around the epithelial cell. Training was broadly performed using ipsilateral, contralateral, chest wall, DCIS and metastatic recurrences. Cross-validation findings were strong for both classifiers (Fig. 3A, B). As phospho-Ser-226-GLUT1 and PFKL are related to different signaling events (6), monitoring both biomarkers should miss fewer positive samples. Previous in vitro studies report that exosomes and lncRNA transfer the cancerous metabolic phenotype to other cells (7). Such intercellular trafficking events may contribute to low sampling error. In unpublished studies we found that 75% of atypical ductal hyperplasia (ADH) recurrences were predicted by the phospho-Ser226-GLUT1 classifier (Schanen et al., unpublished). The decision to recur may take place during ADH (a lesion likely preceding DCIS), thereby reducing uncertainty in DCIS tests.

Our ML models show the presence of FPs in raw patient data (Table 1). This was expected because 98% of the patients tested underwent full or partial mastectomies, and surgeries are expected to cure some patients. Indeed, FPs would be anticipated because the number of recurrences among DCIS patients treated with biopsy alone (47 ± 21%) is much greater than the observed number of recurrences after partial mastectomy (10%) (20-29, 33). In these populations, one would infer that surgery cures many patients who would otherwise recur. Nonetheless, how do we know that this is true for the cohort of patients reported herein? Six lines of reasoning support this proposal. 1) We have used 185 patients in this study, which indicates that the means are very well-known. 2) The underlying glycolytic transporter/enzyme patterns in FPs and TPs are indistinguishable (Fig. 4A, B). 3) Images of recurrent and non-recurrent DCIS tissue were mathematically deconstructed and patterns unique to recurrent and non-recurrent samples were found that did not distinguish between TP and FP. For clinical recurrences, the two classifiers in tandem perform without error. It is likely that the classifiers perform equally well in the analysis of FP micrographs. 4) Moreover, a cutline of 98% suggests that FP lesions contain robust recurrent tissue. 5) The percentage of recurrence positive microscope fields in TP lesions is indistinguishable from that of FP lesions, which suggests that the nature of FP lesions cannot be explained quantitatively by the number of positive microscope fields. 6) The properties of FP in the present study are also supported by ongoing studies of ADH, a lesion similar to DCIS that was not treated with surgery. When evaluated with the phospho-Ser-226-GLUT1 ML model, the FP were reduced to only 10% of the TN population in comparison to ∼50% of the TN for surgery-treated DCIS. Thus, population-based data in the literature and our ML analyses suggest that full and partial mastectomies contribute to the FPs observed. However, it remains possible that some of FPs were TNs because a downstream step in cancer aggressiveness was not functional, and thereby blocked cancer recurrences.

To our knowledge, this is the first prognostic test designed to perform well using samples from African American women. We underscore the need to build machine classifiers without racial bias. African American women must be included in computer training and patient holdout groups. Machine classifiers should be constructed such that morphological features correlating with race, but not cancer recurrences or non-recurrences, are present in both training data sets. For example, reduced blood vessel elasticity in African Americans (39) likely impacts machine predictions. By balancing sample features in machine training, outcome predictions are improved.

Prognostic tests must display clinical utility and patient benefits (40). Clinical utility is supported by the fact that patient outcomes will be improved. For recurrent patients, this test will reduce the uncertainty of cancer recurrence and help plan treatments. If ML findings show that normal adjacent tissue is positive for glycolytic activation patterns (6), a full mastectomy should be considered. Patients predicted to be non-recurrent will benefit from a reduction in the unnecessary burdens of surgery, financial costs, disfigurement, pain, sensory disturbances, and psychological damage (41-43). It is also possible that radiotherapy could cause secondary tumors in normal breast tissue. Instead, negative patients could be safely monitored by mammography. This tool will alert physicians to future recurrences and reduce the impact of overdiagnosis.

In addition to patient management, this work contributes to our understanding of the Warburg Effect. Although the Warburg Effect has been studied for many years, our fundamental understanding of this effect has lagged. The Warburg Effect may exist because it produces ATP faster than respiration (44), although it is much less efficient. Nonetheless, we still do not clearly understand how the system up-regulates glucose transport. This work provides insight into the biochemical mechanism of Warburg’s Effect. We suggest that trafficking of phospho-Ser-226-GLUT1 and PFKL from the nucleus to the cell periphery in recurrent (or aggressive) cancer increases the rate of glucose equilibration across the epithelial cell surface and the formation of PFKL’s products, which is accomplished with reduced feedback inhibition. As phospho-Ser-226-GLUT1 and PFKL mobilization appear to be early steps in the recurrence mechanism, stabilization of nucleoli would seem be a potentially useful route in drug discovery.

ML will become an <u>in-silico</u> research tool. As TP and FP classes contain computed recurrent and non-recurrent microscopic fields, it may be difficult to characterize these lesions with non-microscopic tests. Genomic changes in DCIS recurrences may be identified by combining our machine tool with single cell genomics (45). Our ML models in conjunction with composite DCIS micrographs containing image features from both recurrent and non-recurrent samples suggest that recurrences are promoted by phospho-Ser226-GLUT1 of epithelial cells, blood vessels, and cancer associated fibroblasts (8), as judged by the abilities of these image features to convert non-recurrence into recurrence-positive images (8). Computed saliency maps will also permit identification of cells and organelles contributing to cancer recurrences. Thus, we may now ask more detailed questions about the mechanism of breast cancer recurrences, and it may become possible to better control patient outcomes. There is little doubt that ML will promote new approaches in cancer research, improve healthcare, and expand personalized medicine.

## Conclusions

Our ML models identify women at a low risk for cancer recurrence and women at a high risk of recurrence whose tissues display patterns of glycolytic elements that correlate with enhanced glycolytic activity. In addition to providing insights and a prognostic test relevant to the long-standing problem of overdiagnosis, these data support a biophysical/biochemical mechanism of breast cancer recurrences that may contribute to a new generation of drugs that are specifically directed against cancer recurrences.

## Data Availability

All aggregated data produced in the present study are available upon reasonable request to authors.

## Acknowledgements

We thank the St. Louis Breast Tissue Registry at Washington University School of Medicine and Beaumont Hospital for assistance in obtaining tissue samples. We thank Dr. L.I. Petty for editing this manuscript. This work was supported by funding from the Mildred E. Swanson Foundation and the Michigan Economic Development Corp.

---

**Supplementary Figure 1.**
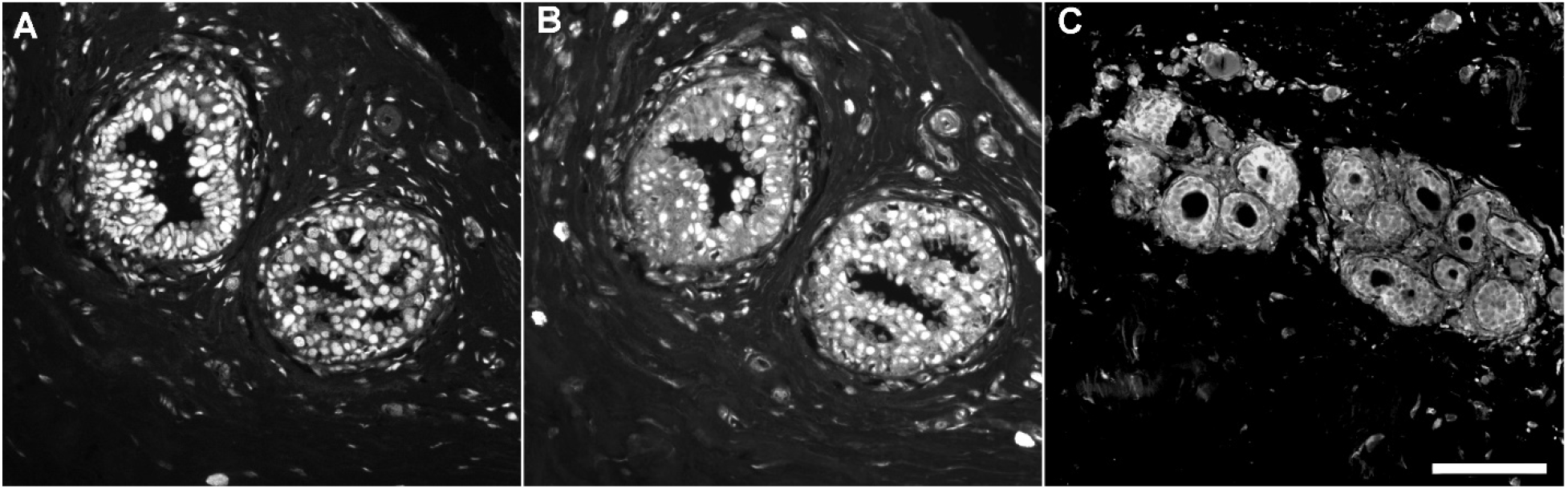
An example of a non-representative DCIS case that was clinically non-recurrent and computationally non-recurrent. Panel A shows a DCIS lesion stained with anti-H2A.X antibodies. Panel B shows the same sample stained with anti-phospho-Ser226-GLUT1. Panel C shows anti-PFKL labeling of this patient sample. As H2A.X is a nucleolar marker, panels A and B suggest that phospho-Ser226-GLUT1 is in the vicinity of the nucleus in DCIS lesions of non-recurrent patients. The central location of phospho-Ser226-GLUT1 has been previously noted (5-9). On the other hand, PFKL was found at the periphery of ductal epithelial cells (panel C). Although peripheral PFKL is consistent with a cancer recurrence (6), All phospho-Ser226-GLUT1 and PFKL micrographs (>200) were computationally negative. This suggests that computer vision is a more sensitive detector of cancer recurrences than human inspection of micrographs.

